# Reduced T cell and antibody responses to inactivated coronavirus vaccine among males and individuals above 55 years old

**DOI:** 10.1101/2021.08.16.21262069

**Authors:** G.X. Medeiros, G.L. Sasahara, J.Y Magawa, JPS Nunes, F.R. Bruno, A. Kuramoto, R.R. Almeida, M.A. Ferreira, G.P. Scagion, E.D. Candido, F.B. Leal, D.B.L. Oliveira, E.L. Durigon, R.C.V. Silva, D.S. Rosa, S.B. Boscardin, V.P.C. Coelho, J Kalil, K.S. Santos, E. Cunha-Neto

## Abstract

**Background:** CoronaVac is an inactivated SARS-CoV-2 vaccine that has been rolled out in several low and middle-income countries including Brazil, where it was the mainstay of the first wave of immunization of health care workers and the elderly population. We aimed to assess the T cell and antibody responses of vaccinees as compared to convalescent subjects.

**Methods:** We detected IgG against SARS-CoV-2 antigens, neutralizing antibodies against the original SARS-CoV-2 strain, and used SARS-CoV-2 peptides to detect IFN-g and IL-2 specific T cell responses in a cohort of CoronaVac vaccinees (N=101) and convalescent (N=72) individuals.

**Findings:** Among vaccinees, 95% displayed T cell or antibody responses to SARS-CoV-2 as compared to 99% convalescent individuals. However, we observed that among vaccinees, males and individuals 55 years or older developed significantly lower anti-RBD, anti-NP and neutralizing antibody responses as well as antigen-induced IL-2 production by T cells.

**Interpretation:** Even though some studies indicated Coronavac helped reduce mortality among elderly people, considering the current dominance of the gamma variant of concern (VOC) and potential increase of the delta VOC, in Brazil, our data support that Coronavac vaccinees above 55 years old Coronavac vaccinees above 55 years old could benefit from a heterologous third dose/booster vaccine to improve immune response and protection.

**Funding:** Brazilian Ministry for Science, Technology and Innovation, Sao Paulo State Foundation for Scientific research (FAPESP), JBS S.A.

## Background

Terminating the COVID-19 pandemic is dependent on global vaccination. CoronaVac (Sinovac, China) is a vaccine based on inactivated SARS-CoV-2 that has been deployed in China, Brazil, Indonesia, Thailand, Turkey, and Chile among other countries. In Brazil, Coronavac was the mainstay of the first wave of immunization of health care workers and the elderly population. In spite of the finding of reduced COVID-19 mortality in Brazil among people above 70 or 75 years of age were registered when Coronavac was the most used vaccine, indicating protection for this group, immunogenicity of this vaccine in elderly individuals is still poorly known.^1,2^ Some studies reported seroconversion for up to 98% of vaccinees but anti-Spike antibody titers were significantly lower among those aged ≥ 60 years.^3,4^ The immunogenicity of inactivated vaccines such as influenza is more limited among the elderly.^5,6^

MRNA-based vaccines that protect more than 90% of the vaccinated individuals from severe COVID-19 were shown to induce T cell responses.^7^ Although an immunogenicity study in Chile has studied cellular immunity to Coronavac, few subjects were above 60 years of age.^8^ In order to assess the effect of age and sex in the vaccine response of adults and elderly people, we studied the anti-SARS-CoV-2 responses of a group of 101 vaccinees (including 42 subjects above 60) as compared to COVID-19 convalescent and seronegative individuals. In this paper, we assessed T cell immune responses with an antigen-induced cytokine release assay on whole blood as well as both binding antibody responses (against Spike, RBD and NP) and neutralizing antibodies against the original Wuhan strain.

## Methods

Coronavac vaccinees (n=101; age range 23-90) received two doses of 3 ug vaccine/shot, 3 weeks apart. Venous blood was collected at least 28 days after the second immunization. Convalescent individuals (n=72; age range 25-68) with at least 160 days since the onset of the infectious episode, as well as seronegative controls (n=36; age range 22-66) were also studied. All volunteers signed written informed consent and the study was approved by the Ethics Committee of the Hospital das Clinicas da Universidade de São Paulo (CAPPesq CAAE30155220.3.0000.0068).

Enzyme-linked immunosorbent assay (ELISA) was performed using 96-well high-binding half-area polystyrene plates coated overnight at 4°C with 4 μg/mL of Spike protein, 2 μg/mL Nucleocapsid protein (NP) (Kindly provided by Dr Ricardo Gazzinelli, UFMG, Brazil) or 0·8μg/mL of the RBD domain from SARS-CoV-2 were all expressed in HEK293T cells.^9^ In short, 50 µL of diluted sera (1:100) were incubated at 37°C for 45 min. Peroxidase-conjugated goat anti-human IgG (BD Pharmingen,USA), anti-human IgA (KPL, USA) or anti-human IgM (Sigma, USA) secondary antibody conjugates were diluted 1:10,000, and incubated at 37°C for 30 min. Values were determined as optical density (OD) minus blank and cutoff was determined as blank+ 3x standard deviation. Results are given as the ratio of OD sample/cutoff). An antibody ratio of ≥1·2 was considered positive.

SARS-CoV-2 (GenBank: MT MT350282) was used to conduct a cytopathic effect (CPE)-based virus neutralization test (VNT) as previously described.^10^ We used 96-well plates containing 5 × 10^4^ cells/mL of Vero cells (ATCC CCL-81). A series of dilutions (1:20 to 1:2560) was prepared for the assay. Serum dilutions were mixed at equal volumes with the virus (100 tissue culture infectious doses, 100% endpoint per well – VNT_100_) and pre-incubated for virus neutralization for 1 hour at 37°C. The mixtures containing serum and virus were transferred onto the confluent cell monolayer and incubated at 5% CO_2_ for three days at 37°C, all the procedures were conducted in a Biosafety laboratory level three (BSL3). After 72 hours, plates were analyzed by light microscopy. Gross CPE was observed on Vero cells, distinguishing the presence/absence of CPE-VNT. To determine neutralizing antibody titers, the highest serum dilution that was able to neutralize virus growth was considered. As positive control, a reference serum from a RT-qPCR positive individual and a plaque reduction in the neutralization test >640 was used in each assay. Antigen-induced T cell cytokine release assays were performed by incubating 250 µl/well of heparinized peripheral blood onto round-bottom 96 well plates for 48h at a humidified, 37°C, 5% CO2 environment in the presence of 20 pooled CD4+ T cell epitopes. Blood samples were kept at 4°C for 16h, and rewarmed at 37°C for 1 h, prior to plating. The 20 CD4+ T cell epitopes used as antigen-specific T cell stimulus were bioinformatically identified and synthesized by scanning the whole proteome in SARS-CoV-2 reference genome (RefSeq: NC_045512.2) using the promiscuous HLA-DR binding peptide approach.^11^ Plates were centrifuged and plasma supernatant was harvested and stored at -80°C until use. IFN-γ and IL-2 levels in cell-free culture supernatants were evaluated by ELISA, according to manufacturer’s instructions (R&D Systems, Minneapolis, MN). Cytokine values were subtracted from the DMSO control. The quantification limit of the IFN-γ test was 1·17 pg/mL and 0·98 pg/mL for IL-2.

Statistical analyses were performed using GraphPad Prism version 9 and a p-value < 0·05 was considered statistically significant. Continuous variables were analyzed using Kruskal-Wallis test with Dunn’s post-hoc test for multiple comparisons when three or more groups were compared, or Mann-Whitney test when two groups were compared. Feature age was clustered using the non-hierarchical K-means algorithm considering humoral or cellular responses.

## Results

The subject groups and assays are described in Supplemental Table I. Among vaccinees, 95% displayed antigen-induced cellular cytokine and/or antibody responses to at least one antigen tested, while 99% of convalescent subjects displayed cellular or antibody responses. Of interest, subjects displaying both cellular and humoral responses were 59 and 70%, among vaccinees and convalescent individuals respectively (Fig. 1A). Vaccinated and convalescent individuals displayed significantly higher antibody and T cell responses than unexposed individuals. Vaccinated individuals displayed significantly reduced IgG responses against Spike protein, but increased responses against the RBD domain as compared with convalescent subjects, while responses to NP were similar between those two groups (Fig. 1B).

**Figure 1.**
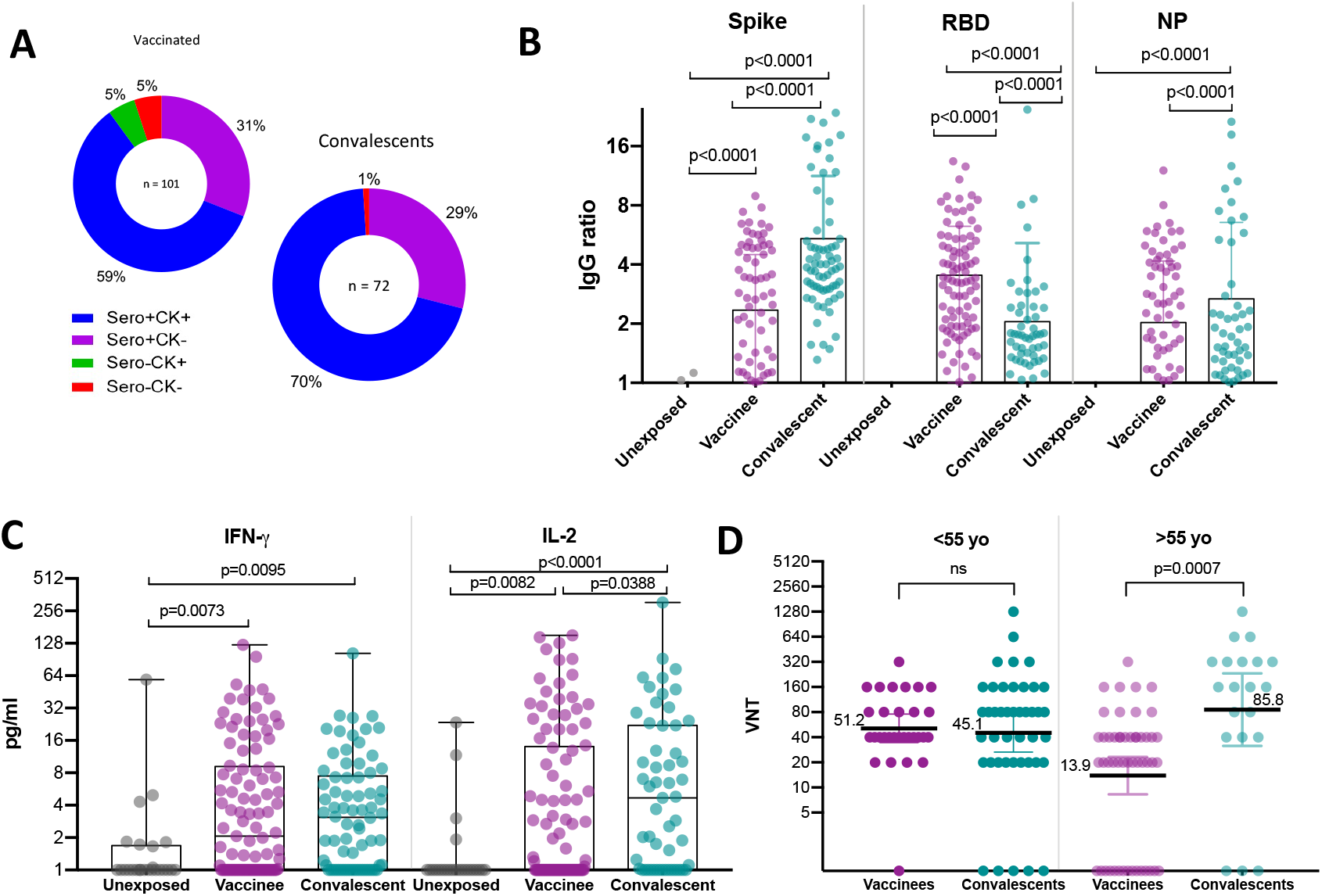
Immune reponses comparing vaccinees and convalescents. A: IgG and T-cell-dependent SARS-CoV-2-specific cytokine production among vaccinees and convalescent subjects. B: IgG reactivity against SARS-COV-2 Spike protein, RBD domain and nucleocapsid protein C: T-cell-dependent SARS-CoV-2-specific cytokine release upon whole blood stimulation with specific SARS-COV-2 peptides. D: Viral neutralization titers grouped by age. VNT: Virus Neutralization Titer; NP: Nucleocapsid Protein from SARS-COV-2; IFN-g: Interferon gamma. VNT below 1:20 were considered 1 in graphs, numbers above the bars show the Geometric Mean Titer (GMT), and the error bars indicate the 95% CI.

Regarding T cell responses as measured by cytokine release after peptide stimulation, we observed that IL-2 levels were higher among the convalescent as compared to the vaccinated group, and both were higher than the unexposed group, while IFN-gamma levels were similarly increased among vaccinees and convalescent subjects (Fig. 1C). GMT of neutralization titers for the 55 and older group were 6 times lower among vaccinees as compared to convalescent patients, while there was no significant difference between convalescent and vaccinated groups <55 yo. (Fig. 1D).

Most immune response levels were positively correlated among each other, as previously reported,^12^ except for NP and Spike (Fig. 2). Neutralization titers positively correlated with IgG levels for the three antigens tested and also with IFN-g and IL-2 production upon whole blood stimulation (Fig. 3). Among vaccinees, we observed significant negative correlations between age and anti-Spike, anti-RBD and anti-NP IgG antibody levels as well as IL-2 cytokine release (Fig. 4). The identification of clusters from the numerical variables allowed us to identify the age of ∼55 years as the best divisor. Therefore, we compared the two age groups: under 55 with 55 and older.

**Figure. 2.**
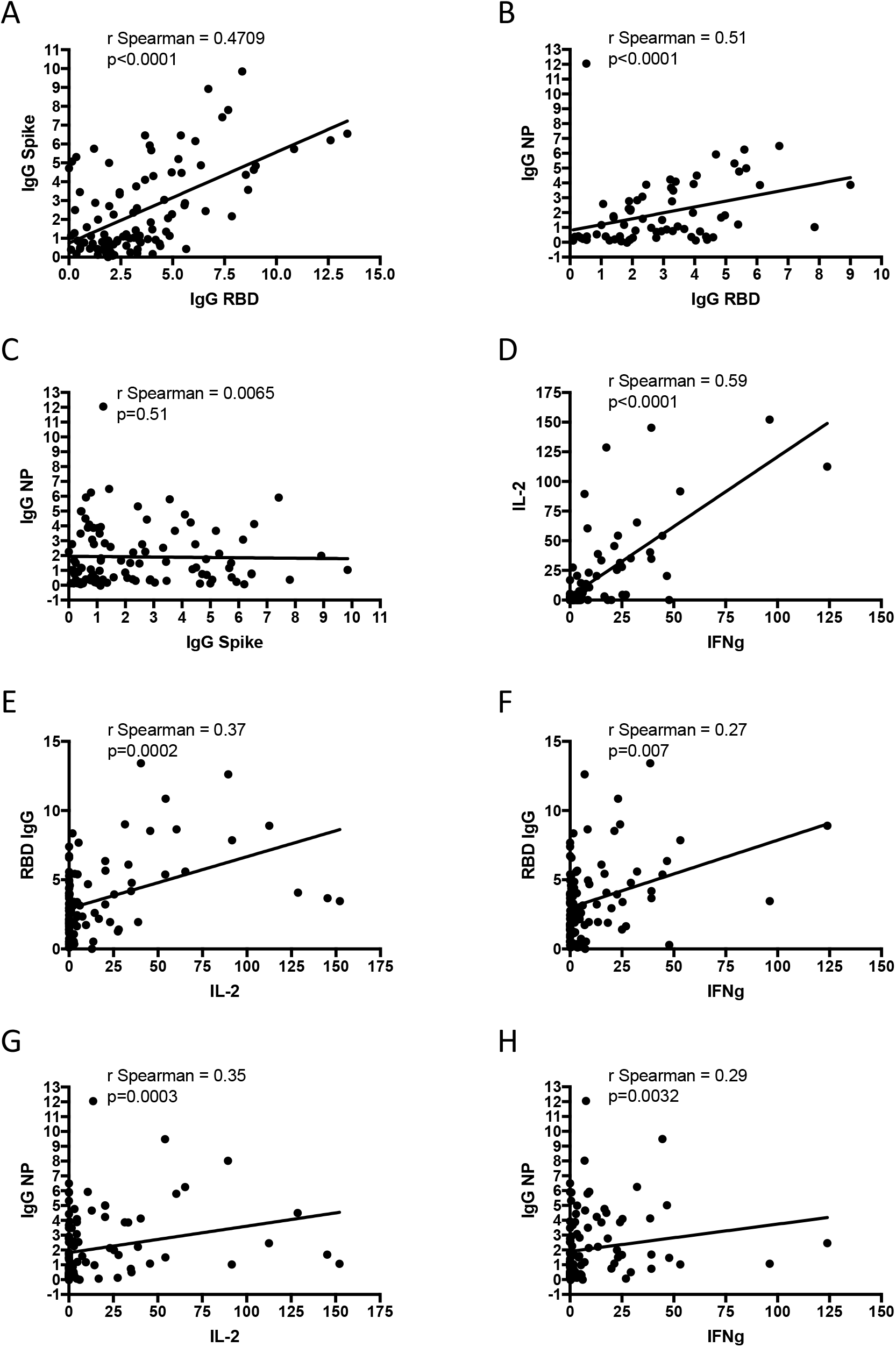
Correlations between diferente immunological parameters among vaccinees. A: IgG for Spike protein and RBD B: IgG for NP protein and RBD C: IgG for NP protein and Spike protein. D: IL-2 and IFN-g released after whole blood stimulation E: IgG for RBD and IL-2 released after whole blood stimulation F: IgG for RBD and IFN-g released after whole blood stimulation G: IgG for NP and IL-2 released after whole blood stimulation H: IgG for NP and IFN-g released after whole blood stimulation NP: Nucleocapsid Protein from SARS-COV-2; IFN-g: Interferon gamma.

**Figure. 3.**
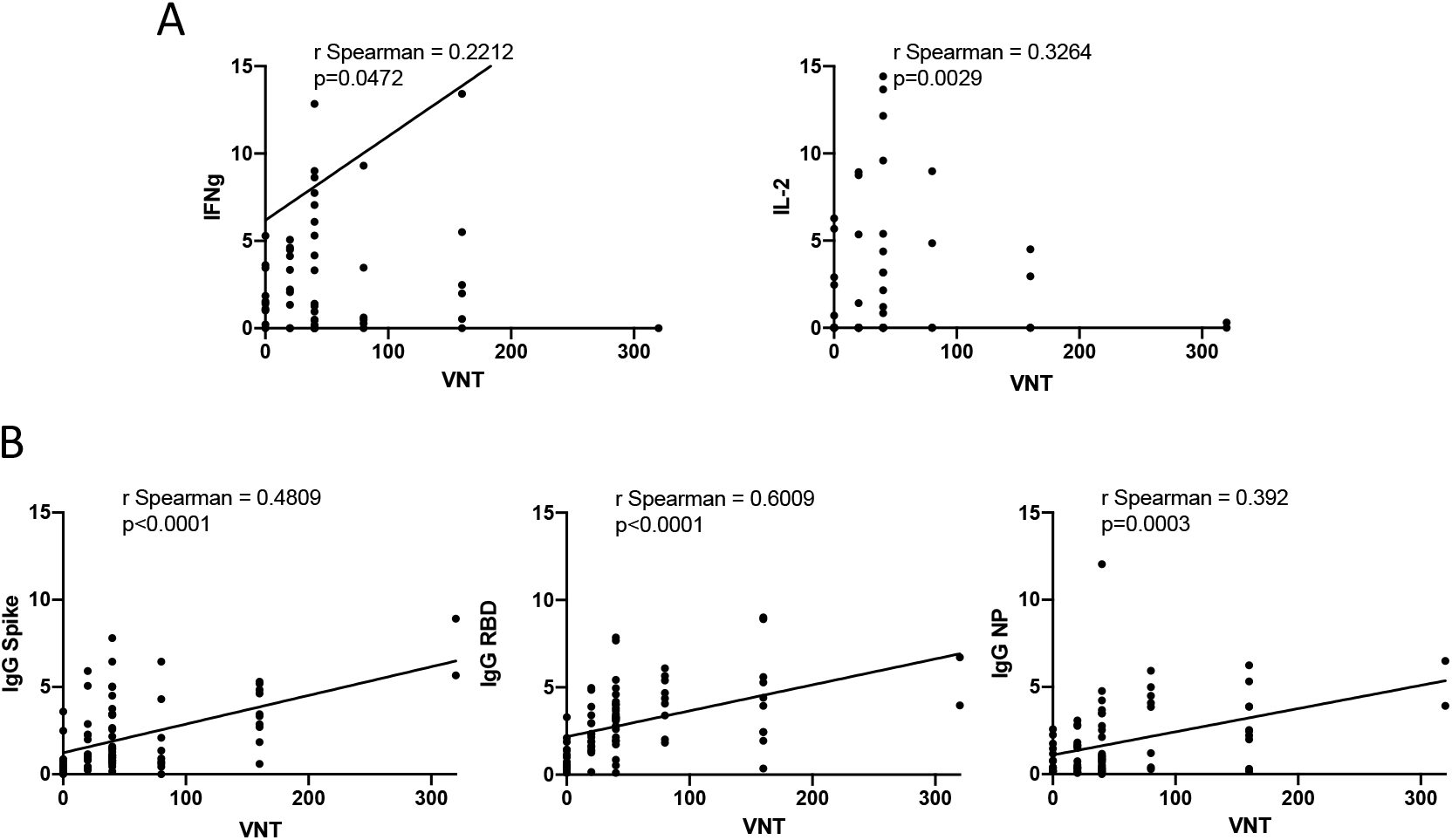
Correlations between Viral Neutralization Titers and other immunological parameters among vaccinees. A: VNT and T-cell responses: IFNg and IL-2 released after whole blood stimulation. B: VNT and humoral responses: IgG for Spike, RBD or NP protein. VNT: Virus Neutralization Titer; NP: Nucleocapsid Protein from SARS-COV-2; IFN-g: Interferon gamma. VNT below1:20 were considered 1 in graphs.

**Figure. 4.**
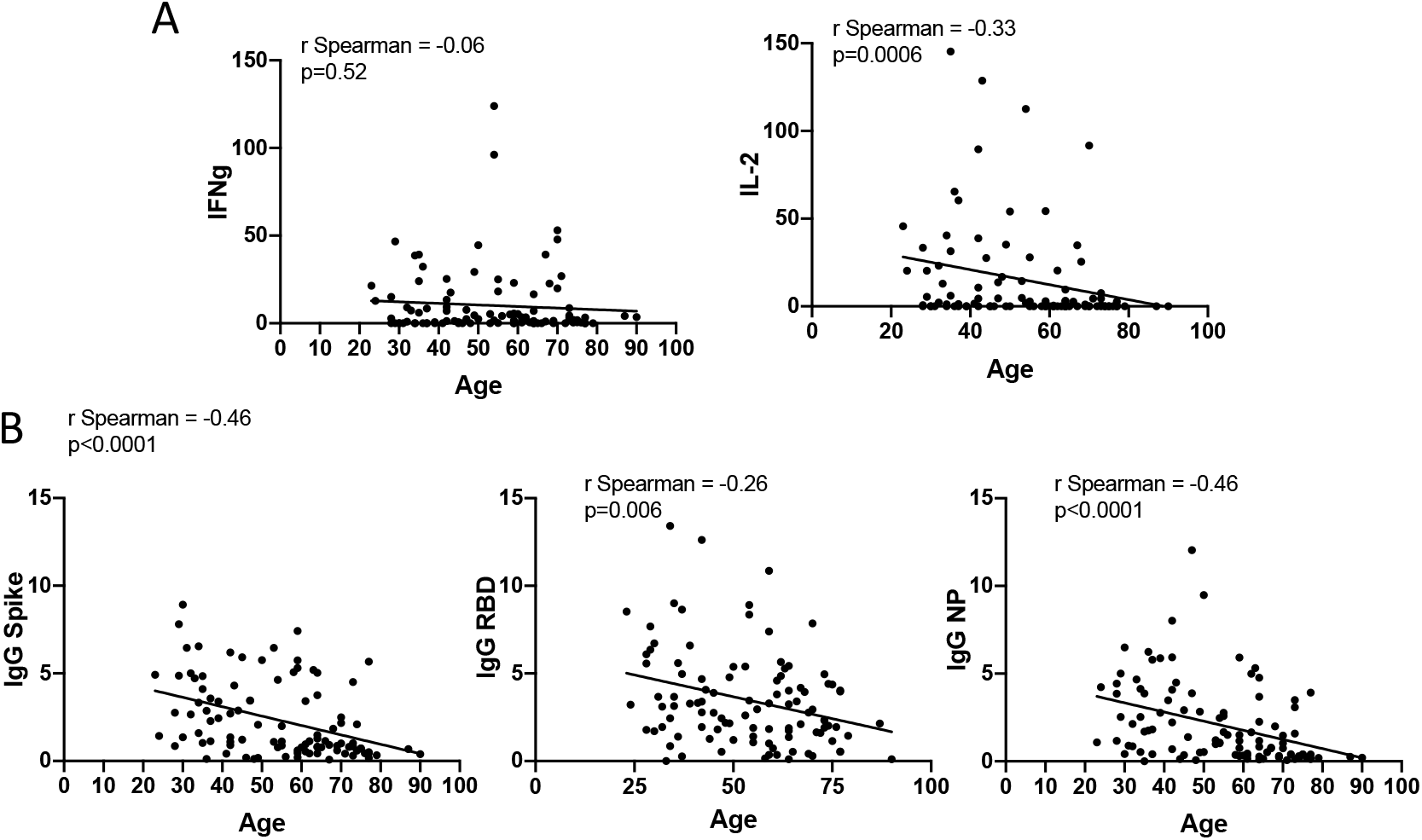
Correlations between age and other immunological parameters among vaccinees. A: Age and T-cell responses: IFNg and IL-2 released after whole blood stimulation. B: Age and humoral responses: IgG for Spike, RBD or NP protein. NP: Nucleocapsid Protein from SARS-COV-2; IFN-g: Interferon gamma.

While 94% of the female vaccinees ≥55 displayed antibody or T cell responses, only 83% of male subjects displayed any detectable responses (p<0·01, Fisher’s exact test). Moreover, whilst 60% of females ≥55 displayed antibody and T cell responses, only 28% of men in the same age group presented both types of response simultaneously (p<0·01, Fisher’s exact test). Antibody responses alone were observed in 39% vs 31% of male and female vaccinees ≥55 (p<0·001), respectively, and cellular responses in the absence of detectable IgG was found among 17% male and 3% female vaccinees ≥55 years old (Fig. 5A). Similar results were observed when we assessed neutralizing antibody and T cell cytokine production (Fig. 6).

**Figure 5.**
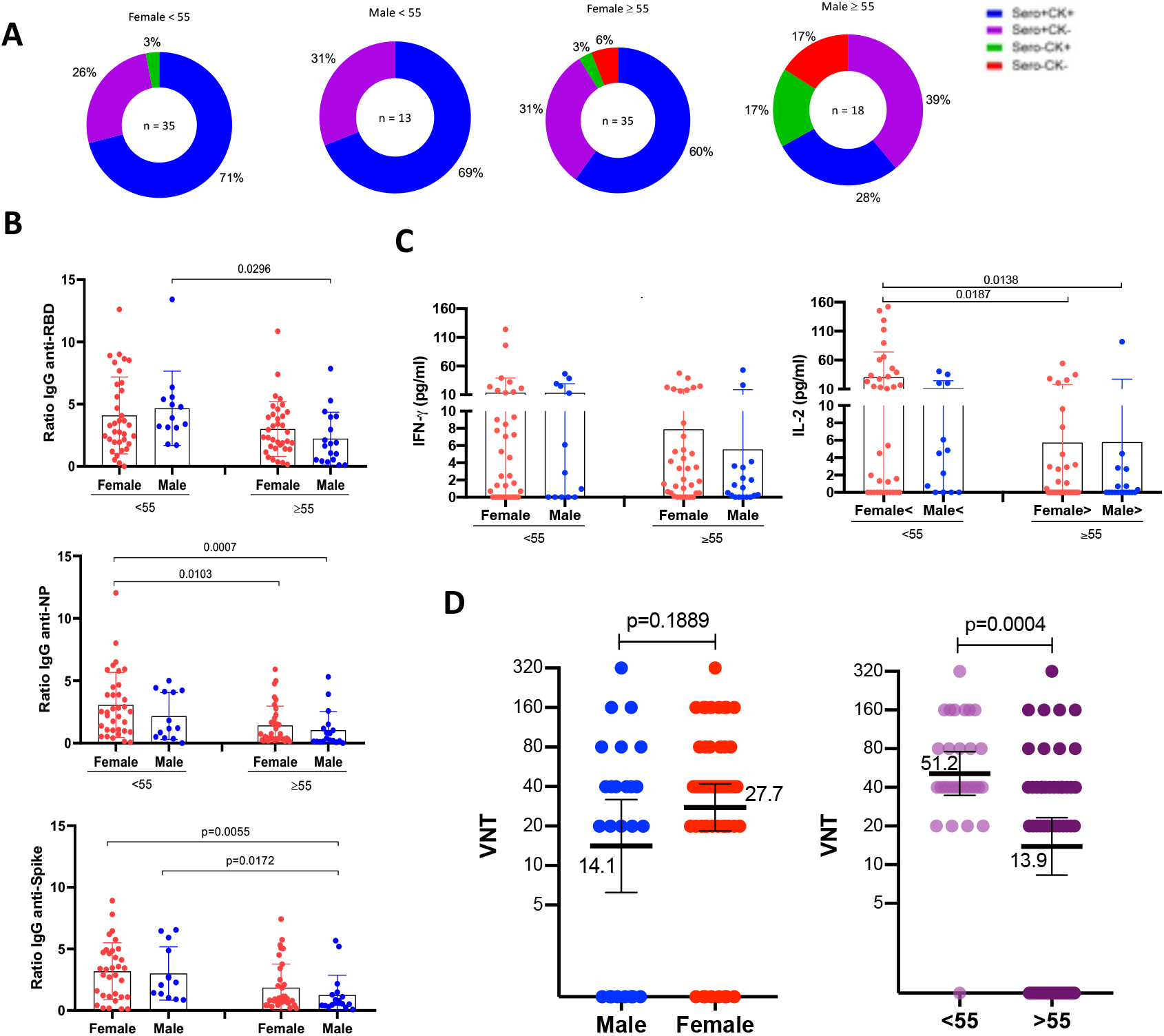
Immune responses among Coronavac vaccinees. A: IgG and T-cell-dependent SARS-CoV-2-specific cytokine production among vaccinees. (n=101). B: IgG reactivity against SARS-COV-2 Spike protein, RBD domain and nucleocapsid protein and T-cell-dependent SARS-CoV-2-specific cytokine release upon whole blood stimulation with specific SARS-COV-2 peptides grouped by age C: Viral neutralization titers among vaccinees grouped by age and sex (n=81). VNT: Virus Neutralization Titer; NP: Nucleocapsid Protein from SARS-COV-2; IFN-g: Interferon gamma. VNT below 1:20 were considered 1 in graphs, numbers above the bars show the Geometric Mean Titer (GMT), and the error bars indicate the 95% CI.

**Figure. 6.**
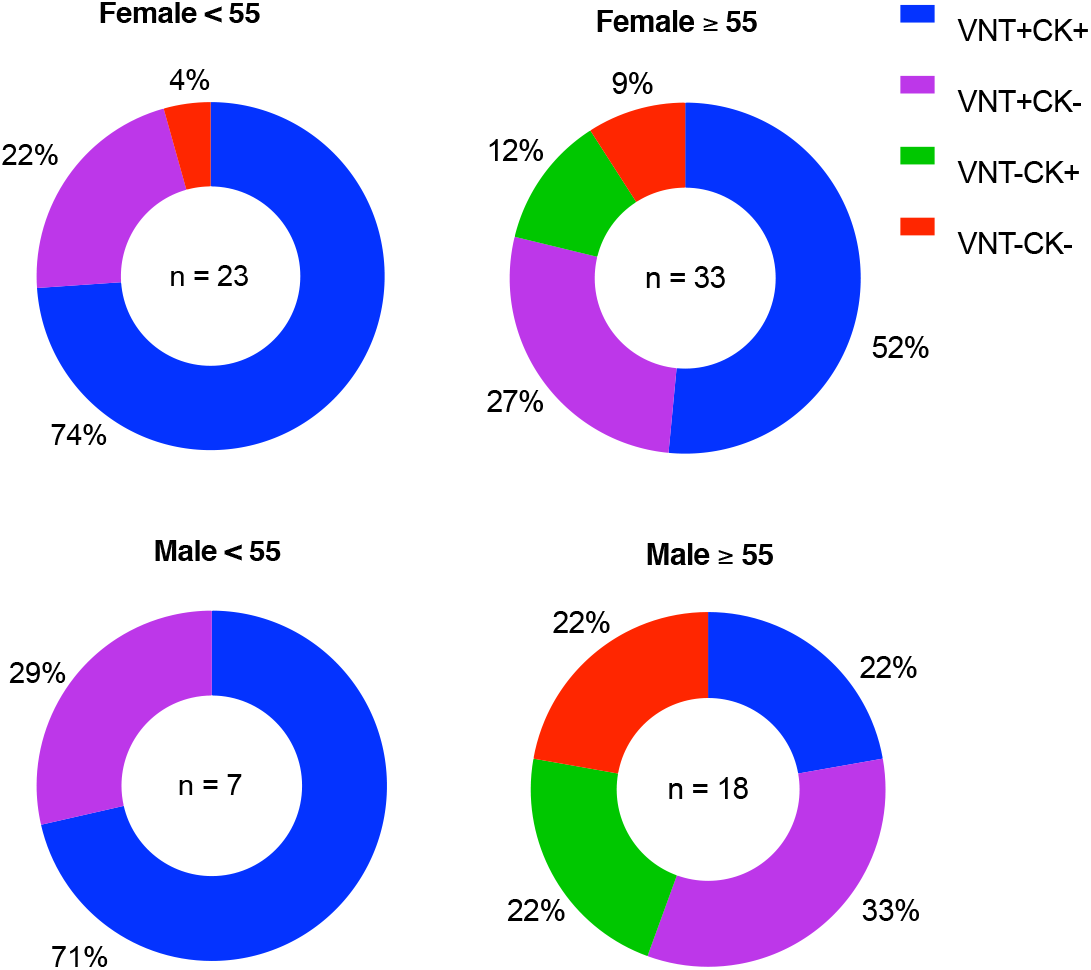
Frequency of immune responses of vaccinees considering VNT titers. Distribution of VNT and/or CK responses among vaccinees.

Antibody responses for male vaccinees ≥55 years old displayed the lowest levels of anti-Spike, anti-RBD and anti-NP IgG and IL-2 release. Female vaccinees ≥55 years old also showed lower anti-RBD IgG and IL-2 release than younger females (Fig. 5B). Likewise, we observed 3·6-fold lower GMT neutralization titers in the vaccinee group ≥55 years old as compared with the younger group (Fig. 5C). GMT neutralization titers against the Wuhan strain were 3-fold lower for vaccinees ≥55 than the younger vaccinees.

## Discussion

Coronavac was the first anti-COVID vaccine available for mass immunization in Brazil, where it followed schedules targeting first the healthcare workers and the elderly population. In our study, the majority of studied vaccinees developed some kind of immune response to the vaccine and the magnitude of most of the immune response measures positively correlated with each other. However, we observed a negative correlation between age and SARS-CoV-2-peptide epitopes antigen-induced IL-2 release as well as antibody responses to Spike, RBD and NP, suggesting and immunogenicity decrease in older individuals. Upon stratification by age and sex, we observed that most of this reduced immunogenicity was due to the ≥55 yo male population. Specifically, the proportion of individuals who displayed any response to the vaccine varied from 100% among younger females (<55 years old) to 83% among males 55 or older. Significantly, the simultaneous detection of antibody and cellular immune responses, the ideal situation, varied from 71% among females younger than 55 years old to only 28% among males 55 or older. This suggests that the immunogenicity of the vaccine regimen is less pronounced in this age and sex group. Indeed, interleukin-2 is produced by central memory T cells and is essential for the development of T cell memory.^13^

COVID-19 immunogenicity across young and elderly patients has been studied for other vaccines. For ChadOx1, no differences were found following the second dose either for anti-Spike IgG and neutralizing antibodies or for IFN-γ and IL-2 Th1 T cell responses among the 18-55, 55-69 and ≥70 groups.^14,15^ A study between mRNA-1273 vaccine recipient groups of 56-70 or ≥71 years of age revealed that binding, neutralizing-antibody and IFN-γ and IL-2 responses were similar to those previously reported among vaccine recipients between the ages of 18 and 55 years and were above the median of a panel of controls who had donated convalescent serum.^7^ In another study, responses against the Pfizer BNT162b1 among Chinese subjects aged 18-55 and 65-85 years of age showed similar IgG and somewhat lower neutralizing antibody and a more variable T cell response than younger individuals.^16^ T cell and antibody immune responses of the elderly groups after mRNA or adenovirus vector vaccines were thus found to be largely similar to those of younger individuals, which is in contrast with observations in our cohort of inactivated virus-based Coronavac vaccinees. In our study, we used a whole blood IFN-γ and IL-2 release assay, while the cited studies used ELISPOT and flow cytometry analysis, which in theory could be more sensitive than the whole blood cytokine release assay used here. However, IgG antibody and VNT assays were similar to those used in the previous studies and showed lower percentages among elderly individuals - especially in the male sex. This is especially relevant given the finding that Coronavac vaccinees have diminished significantly reduced neutralizing capacity against all VOCs alpha, beta and delta.^17^

In summary, our results show a diminished overall immune response for people older than 55 years after two-dose immunization with inactivated vaccine Coronavac. Given the finding that mixing vaccines with different platforms elicits stronger immunogenicity, our results may suggest the Coronavac vaccinees above 55 years old could benefit from a heterologous third-dose/booster vaccine.^18^

**Table 1.** Characteristics of the study cohort

| Table 1 \| Characteristics of the study cohort |  |  |  |  |
| --- | --- | --- | --- | --- |
|  | Total | Vaccinees<br>non infected | Convalescent | Unexposed |
| **n** |  | 101 | 72 | 36 |
| **Age in years, mean (s.d.)** |  | 50.73 (14.22) | 40 (10) | 36 (12.3) |
| **Sex, n (%)** |  |  |  |  |
| Female |  | 70 (70.7) | 54 (75) | 26 (72) |
| Male |  | 31 (31.31) | 18 (25) | 10 (28) |
| **Time since dose 2 or**<br>**post symptoms onset,**<br>**mean days (s.d.)** | - | 44 (23) | 215 (63) | - |
| **Antibody levels, mean**<br>**ratio (s.d.)** |  |  |  |  |
| IgG RBD |  | 2.96 (2.73) | 1.46 (3.07) | - |
| IgG NP |  | 1.17 (2.16) | 1.31 (3.85) | - |
| IgG S |  | 1.35 (2.29) | 3.76 (5.84) | - |
| **Neutralization titer, n**<br>**tested; GMT** |  | n=81 ; 1:22 | n=62 ; 1:54 | - |
| **Cytokine levels, mean**<br>**(s.d.)** |  |  |  |  |
| IFN |  | 2.06 (19.53) | 3.39 (13.60) | - |
| IL-2 |  | 1.21 (47.16) | 4.18 (45.61) | - |

GMT: Geometric Mean Titer; s.d.: standard deviation

## Data Availability

all data referred to in the manuscript are available in the manuscript.

## Notes

### Competing Interest Statement

The authors have declared no competing interest.

### Author Declarations

All volunteers signed written informed consent and the study was approved by the Ethics Committee of the Hospital das Clinicas da Universidade de Sao Paulo (CAPPesq CAAE30155220.3.0000.0068).

